# Socioeconomic disparities in subway use and COVID-19 outcomes in New York City

**DOI:** 10.1101/2020.05.28.20115949

**Authors:** Karla Therese L. Sy, Micaela E. Martinez, Benjamin Rader, Laura F. White

## Abstract

**Background:** The United States CDC has reported that racial and ethnic disparities in the COVID-19 pandemic may in part be due to socioeconomic disadvantages that require individuals to continue to work outside their home and a lack of paid sick leave.^1^ However, data-driven analyses of the socioeconomic determinants of COVID-19 burden are still needed. Using data from New York City (NYC), we aimed to determine how socioeconomic factors impact human mobility and COVID-19 burden.

**Methods/Summary:** New York City has a large amount of heterogeneity in socioeconomic status (SES) and demographics among neighborhoods. We used this heterogeneity to conduct a cross-sectional spatial analysis of the associations between human mobility (i.e., subway ridership), sociodemographic factors, and COVID-19 incidence as of April 26, 2020. We also conducted a secondary analysis of NYC boroughs (which are equivalent to counties in the city) to assess the relationship between the decline in subway use and the time it took for each borough to end the exponential growth period of COVID-19 cases.

**Findings:** Areas with lower median income, a greater percentage of individuals who identify as non-white and/or Hispanic/Latino, a greater percentage of essential workers, and a greater percentage of healthcare workers had more subway use during the pandemic. When adjusted for the percent of essential workers, these association do not remain; this suggests essential work is what drives subway use in lower SES zip codes and communities of color. Increased subway use was associated with a higher rate of COVID-19 cases per 100,000 population when adjusted for testing effort (aRR = 1.11; 95% CI: 1.03 – 1.19), but this association was weaker once we adjusted for median income (aRR = 1.06; 95% CI: 1.00 – 1.12). All sociodemographic variables were significantly associated with the rate of positive cases per 100,000 population when adjusting for testing effort (except percent uninsured) and adjusting for both income and testing effort. The risk factor with the strongest association with COVID-19 was the percent of individuals in essential work (aRR = 1.59, 95% CI: 1.36 – 1.86). We found that subway use declined prior to any executive order, and there was an estimated 28-day lag between the onset of reduced subway use and the end of the exponential growth period of SARS-CoV-2 within New York City boroughs.

**Interpretation:** Our results suggest that the ability to stay home during the pandemic has been constrained by SES and work circumstances. Poorer neighborhoods are not afforded the same reductions in mobility as their richer counterparts. Furthermore, lower SES neighborhoods have higher disease burdens, which may be due to inequities in ability to shelter-in-place, and/or due to the plethora of other existing health disparities that increase vulnerability to COVID-19. Furthermore, the extended lag time between the dramatic fall in subway ridership and the end of the exponential growth phase for COVID-19 cases is important for future policy, because it demonstrates that if there is a resurgence, and stay-at-home orders are re-issued, then cities can expect to wait a month before reported cases will plateau.

## INTRODUCTION

As of May 27th 2020, there are over 5 million confirmed cases of COVID-19, the disease caused by SARS-CoV-2 infection, worldwide.^2–4^ Large metropolitan areas in the United States, including NYC, have been hit particularly hard. A number of factors promote transmission in cities, including high population density, which facilitates transmission via close person-to-person contact.^5–8^ In addition to high population density, the contact network of individuals within cities can help supports long chains of sustained disease transmission.^9–11^ The spread of SARS-CoV-2 in big cities like NYC could also be exacerbated by the reliance on public transportation where riders are sometimes tightly packed in confined spaces, physically unable to space appropriately apart for social distancing. Outside of the pandemic period, 45%-51% of NYC residents reported the subway as their primary transportation to work,^12,13^ and over a billion rides are taken annually.^14^

In the United States, state and local governments have introduced various non-pharmaceutical interventions (NPIs) aimed at curbing SARS-CoV-2 transmission. One type of NPI is social distancing, which seeks to increase physical space between individuals by issuing stay-at-home orders, the shut-down of non-essential businesses, and increased working from home. However, these interventions cannot be applied uniformly. Only 25% of workers in the United States are estimated to be able to transition to remote work, meaning there is a continued need for essential workers to leave their home, putting them at increased risk of exposure to SARS-CoV-2 and increasing the likelihood of local transmission within the community. An individuals’ ability to work remotely is contingent on their industry and position, which is frequently associated with socioeconomic status (SES), race, and education level.^15^ There is potential for increased SARS-CoV-2 exposure in low SES communities due to a more limited ability to shelter-in-place, which we term *social distancing inequity*. Importantly, social distancing inequity may further exacerbate existing health disparities, compounding structural inequalities in the US.

Emerging evidence suggest inequities based on race/ethnicity and socioeconomics have put poorer communities and communities of color at higher risk of SARS-CoV-2 infection.^16^ This may be due to a community’s overall ability to stay-at-home or shelter-in-place. Sheltering-in-place may not be feasible or safe for everyone due to vulnerabilities such as food insecurity,^17,18^ unstable housing,^19,20^ or domestic violence.^21,22^ There is, however, limited published data on the intersection of SES and the ability to shelter-in-place. Previous studies of infectious disease transmission have utilized public transportation data as proxy for human mobility and travel patterns.^23–28^ Here we consider subway ridership as a measure of human mobility and the ability of individuals to follow social distancing measures within NYC. We leverage a publicly available database of subway ridership to explore the associations between human mobility, sociodemographic factors, and COVID-19 incidence. Public transportation use during the pandemic may facilitate transmission within the city because it reflects not only close contacts on the subway, but also the amount to which individuals are required to leave their homes for work and other essential activities.

We hypothesize that geographic areas with higher subway use will: 1) be more likely to be socioeconomically disadvantaged, and 2) have greater burden of disease due to social distancing inequity, with a smaller percent of the population afforded the ability to stay home during the pandemic shutdown. We also anticipate that areas that were unable to rapidly decrease subway use were more likely to experience a higher burden of COVID-19. A May 15^th^ analysis by the New York Times reported that 40% or more residents fled the wealthiest neighborhoods of NYC after the pandemic hit; while few residents from lower income neighborhoods left the city.^29^ This indicates that declines in subway ridership likely reflect the ability to either stay-home within NYC or leave the city to less dense residences, both afforded by wealth.

## METHODS

In this study, we used two geographic scales: zip code tabulation areas (ZCTA) and the borough (county) level. Our primary analysis was a cross-sectional analysis conducted at the ZCTA-level using COVID-19 case data reported as of April 26, 2020, and subway data from the week of January 4, 2020 to April 11, 2020. ZCTAs follow census block boundaries; but they are independent of all other statistical and government entities.^6^ ZCTAs provide a balance between the larger county geographic units where spatial heterogeneity would be lost, and the smaller census tracts that might be so small that individuals would often use subway stations outside their census track. New York City comprises five boroughs and 214 zip code tabulation areas. We removed 25 ZCTAs with no population based on the ACS data, because they are associated with individual buildings.^30^ We removed 63 ZCTAs without a subway station; the final analytic dataset for the ZCTA-level analysis included 126 ZCTAs. In addition, we conducted secondary analyses using weekly subway ridership and COVID-19 cases: Bronx, Brooklyn, Manhattan, and Queens. We excluded Staten Island in all analyses, as there are no subway connections from Staten Island to the other boroughs.

### Data

#### Subway data by Neighborhood

Weekly transportation data are collated by the Metropolitan Transportation Authority (MTA) New York City transit, and are publicly available.^31^ The data are comprised of the number of MetroCard swipes made each week for the 472 subway stations in the NYC Subway system, and we aggregated the swipes for each ZCTA. Longitude and latitude of MTA stations were geocoded by the *ggmap* package in R,^32^ and were manually checked for accuracy. The relative change in subway use served as our main measure of human mobility and social distancing. We estimated a standardized change in subway use for each ZCTA during the outbreak (week of March 7, 2020 to April 11, 2020) by subtracting the mean weekly subway use pre-pandemic (week of January 4, 2020 to February 29, 2020) from the number of subway swipes each week during the pandemic and dividing by the pre-pandemic period standard deviation; in short: (*weekly pandemic ridership – weekly pre-pandemic ridership)/(weekly pre-pandemic SD*). Standardizing the ridership by the pre-pandemic standard deviation allows us to view variation in subway ridership, due to the pandemic, relative to natural variation in subway ridership.

#### Subway data by Borough

Subway data were obtained from the Metropolitan Transportation Authority.^31^ Subway use was calculated as the mean of standardized subway use of the individual ZCTAs for each borough.

#### Demographic and socioeconomic status by Neighborhood

We obtained demographic and socioeconomic status information from the most recent 2014–2018 5-year American Community Survey (ACS) at the ZCTA level, published by the United States Census Bureau.^12^ We used the R package *tidycensus* to obtain the data.^33^ We extracted estimates of population size, median individual income, percent of individuals age > 75 years, percent of the population that identifies as Black or African American, Asian, American Indian and Alaska Native, Native Hawaiian and Other Pacific Islander, and/or Hispanic or Latino (i.e., non-white and/or Hispanic/Latino), percent uninsured, percent with a high school education or less, percent non-healthcare essential workers (employed in: *Agriculture, forestry, fishing and hunting, and mining, Construction, Manufacturing, Wholesale trade, Retail trade, Transportation and warehousing and utilities*), and percent healthcare essential workers (occupation: *healthcare practitioners and technical occupations*).

#### COVID-19 data by Neighborhood

We obtained publicly available COVID-19 data from the NYC Department of Health and Mental Hygiene.^34^ Cumulative COVID-19 cases and tests were collated for each ZCTA as of April 26, 2020. We defined the main COVID-19 outcome as the rate of positive COVID-19 cases per 100,000 population. Previous research has shown that testing accessibility is highly variable across the United States,^35^ so we adjusted all models with this outcome by the number of tests done within each ZCTA. This accounts for some of testing accessibility disparities across New York City, to incorporate both case burden and testing capabilities. We also considered two additional outcomes relating to COVID-19: (1) the percentage of positive tests among all tests, and (2) the rate of total tests per 100,000 population.

#### COVID-19 data by Borough

We digitized daily incident case counts and daily tests at the borough-level from March 2, 2020 until April 26, 2020 from the publicly available data from the NYC Department of Health and Mental Hygiene.^34^ We conducted our analysis on the daily time series of incident and cumulative case counts and tests for each of the New York City boroughs. Our main outcome for the borough analysis was log of the cumulative case counts, as this allows us to assess the timing of the exponential growth period. We also describe the time series of the similar outcomes used in the main ZCTA analysis, but at the borough level: (1) the rate of positive COVID-19 cases per 100,000 population, (2) the percentage of positive tests out of the total number of tests, and (3) the rate of total tests per 100,000 population.

### Statistical analyses

#### Neighborhood Subway use

We first conducted descriptive analyses of New York City subway use at the ZCTA level at the onset of the outbreak (i.e. until the week of April 11, 2020). Subsequently, we examined the cross-sectional association between ZCTA-level ACS sociodemographic variables and change in NYC

MTA subway ridership using a linear regression model to assess whether neighborhood-level features were associated with changes in subway use (presumably as a consequence of stay-at-home policies). We hypothesized that the percentage of the population working in essential services may be driving the association between sociodemographic factors and subway use, and thus the confounder for the subway use outcome was the percentage of the population in essential services in each ZCTA.

#### COVID-19 across Neighborhoods

We describe our three main cross-sectional COVID-19 outcomes as of April 26, 2020. We then assessed the association between subway use and sociodemographic variables on COVID-19 outcomes. For the primary outcome, rate of positive COVID-19 tests per 100,000 population, we use a negative binomial generalized linear model with a log link, and the ZCTA population as the offset. We conducted an unadjusted analysis, an analysis adjusted for testing, and an analysis adjusting for testing and confounders. We hypothesized that incomes may be the main confounder of the association between neighborhood-level features and COVID-19; thus, the confounder adjusted for this analysis was median income. The same model was used for the secondary outcome rate of total tests per 100,000 population. The analyses on the percentage of positive tests out of the total number of tests were conducted with a generalized linear model with a binomial distribution and a logit link, and the total number of tests as weights. We ran unadjusted and adjusted analyses for both secondary COVID-19 outcomes. All analyses was conducted in R version 4.0.0.^36^

#### COVID-19 & Subway use across Boroughs

We calculated descriptive statistics on both subway use and COVID-19 outcomes, and conducted Kruskal-Wallis one-way analysis of variance tests to assess whether the rate of SARS-Cov-2, percentage positive among tested, and rate of testing were significantly different between boroughs. A segmented regression was fit to the subway usage data for each borough to estimate the timing of the change in subway usage, or the **“breakpoint”**.^8–10^ We estimated the end of the exponential growth period as the estimated breakpoint in the log of cumulative cases for a segmented regression for each borough. We qualitatively assessed whether the timing of social distancing may have affected the end of the exponential growth period and epidemic doubling time. Epidemic doubling time was calculated using the method previously used in the SARS-CoV-2 outbreak.^37^

## RESULTS

### Characterizing change in human mobility

The mean subway use during the pre-pandemic period was over 25 million swipes per week. Overall, this decreased 69.7% to less than 8 million by the week of April 11, 2020. The timeline and extent of the reduction in subway use after March 4, 2020 varied greatly between the ZCTAs (Supp Figure 1). The mean standardized change in subway use from baseline to April 11 among all ZCTAs was –20.96 (IQR: –25.57 to –16.05). The Bushwick/Bedford-Stuyvesant neighborhood in Brooklyn had the greatest standardized reduction in subway use with a decrease of 42.86, followed by Upper West Side, Manhattan (−40.47). The areas with the least reductions in subway use were Rockway, Queens with standardized decrease of 2.92, and Fort George, Manhattan (−4.02). Figure 1 shows the geographic variability in standardized changes in subway use during the early stages of the pandemic in New York City.

**Figure 1.**
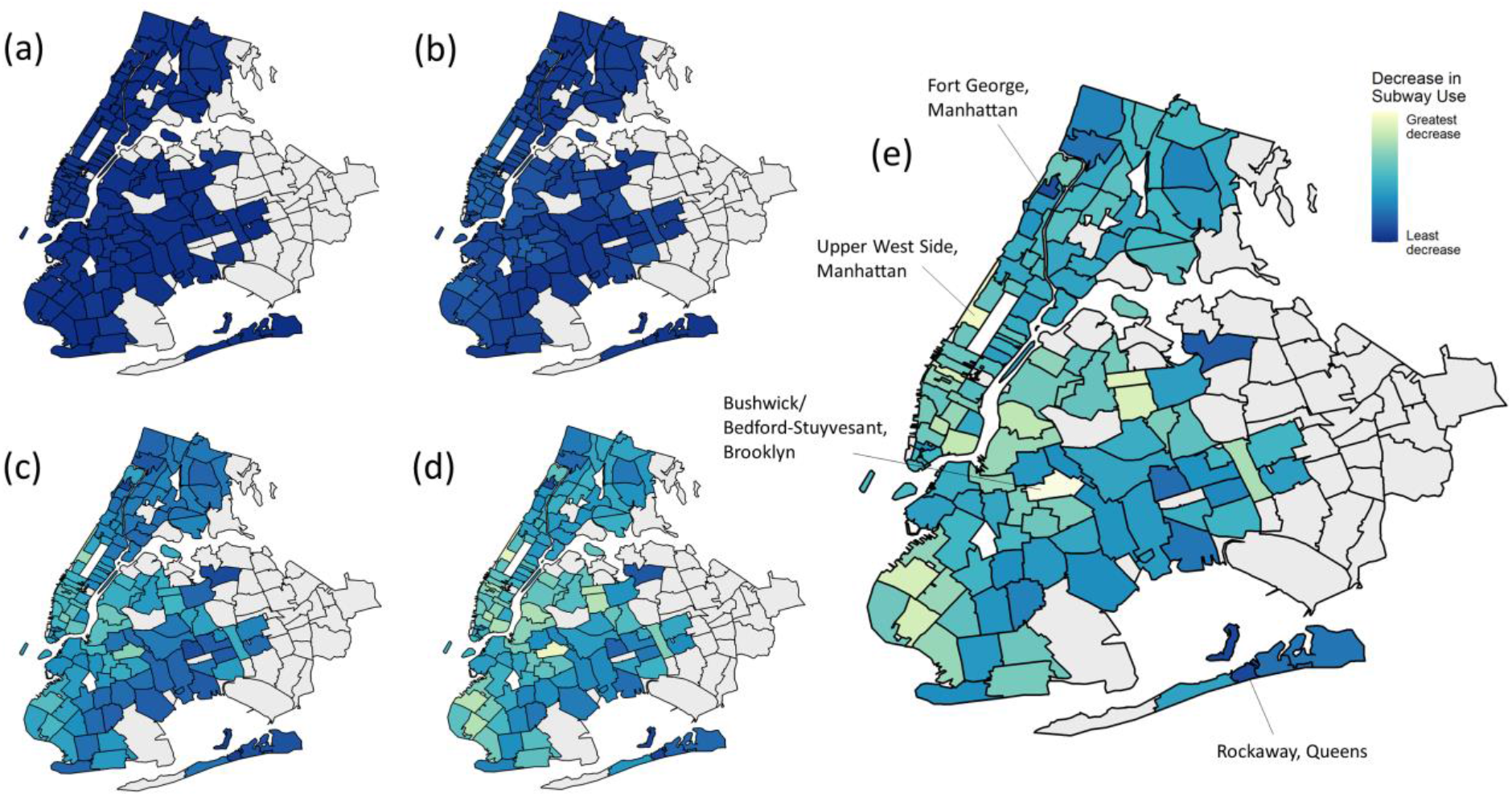
New York City reduction in subway use in ZCTAs during the COVID-19 outbreak on the week of (a) February 29, 2020 (b) March 7, 2020 (c) March 14, 2020 (d) March 21, 2020 (e) April 11, 2020. Reductions are calculated as the change in subway use relative to the prepandemic period and standardized by the pre-pandemic standard deviation. Panels b-d correspond to key New York City executive orders (b) Local state of emergency; restricted gatherings exceeding 500 persons; (c) City schools closure, (d) Stay-at-home order, nonessential businesses closure.^38,54^

The change in subway use also varied between the boroughs (Supp Figure 1). Overall, the mean standardized decrease in subway use was greatest in Manhattan ZCTAs with a decline of 22.4 (95% IQR: 25.65 to –18.44), followed by a reduction of 22.2 (95% CI: –28.17 to –14.72) in Brooklyn, 19.3 (95% CI: –25.08 to –14.72) in Queens and 18.6 (95% CI: –21.36 to –16.76) in the Bronx.

### Characterizing COVID-19 outcomes

Among the ZCTAs as of April 26, the median rate of positive COVID-19 cases per 100,000 population was 2,058.84 (IQR: 1,415.5–2,671.0). The median percentage of cases among tested was 50.98% (IQR: 44.25–55.17), and the median rate of testing per 100,000 population was 4,033 (IQR: 3,135–5,034). The area with the highest rate of COVID-19 (4085.82 cases/100,000 population) and the probability of being COVID-positive given tested (68.67%) was East Elmhurst, Queens. However, the area with the greatest rate of COVID testing was Co-op City in the Bronx with 8262.36 cases per 100,000 population. The areas with the lowest COVID-19 outcomes were all in Manhattan; the area with the lowest rate of COVID (436.91/100,000 population) and lowest testing (1566.50/100,000 population) was Battery Park City in Manhattan, while the area with the lowest percent of cases among those tested was Tribeca, with 26.88%.

By April 26, the Bronx had the greatest rate of COVID-19 among the four boroughs, with 2,472 cases per 100,000 population, followed by Queens (2,149 cases per 100,000 population), Brooklyn (1,653 cases per 100,000 population), and Manhattan (1,292 cases per 100,000 population). Queens had the highest number of cumulative cases among the four boroughs, with 49,399 positive COVID-19 cases. This was followed by Brooklyn with 43,014 cases, the Bronx with 35,556 cases and Manhattan with 21,097 cases. The daily rate of positive cases per 100,000 population (p<0.001), the daily percent of proportion of positives among total tested (p = 0.03), and the daily rate of tests per 100,000 population was significantly different between the four boroughs (p = 0.01) (Supp Figure 2).

## ZIP CODE TABULATION AREA ANALYSIS

### Sociodemographic variables and subway use

The ZCTAs with the highest median income tended to have the greatest decrease in mobility and subway use (Figure 2). In unadjusted analyses, lower median income, larger percent of people working in essential services, larger percent of healthcare workers, and larger percent non-white/Hispanic individuals were associated with a smaller decrease in subway use. In analyses adjusted for percent of people working in essential services, there were no associations with subway use. (Figure 3, Supp Table 1).

**Figure 2.**
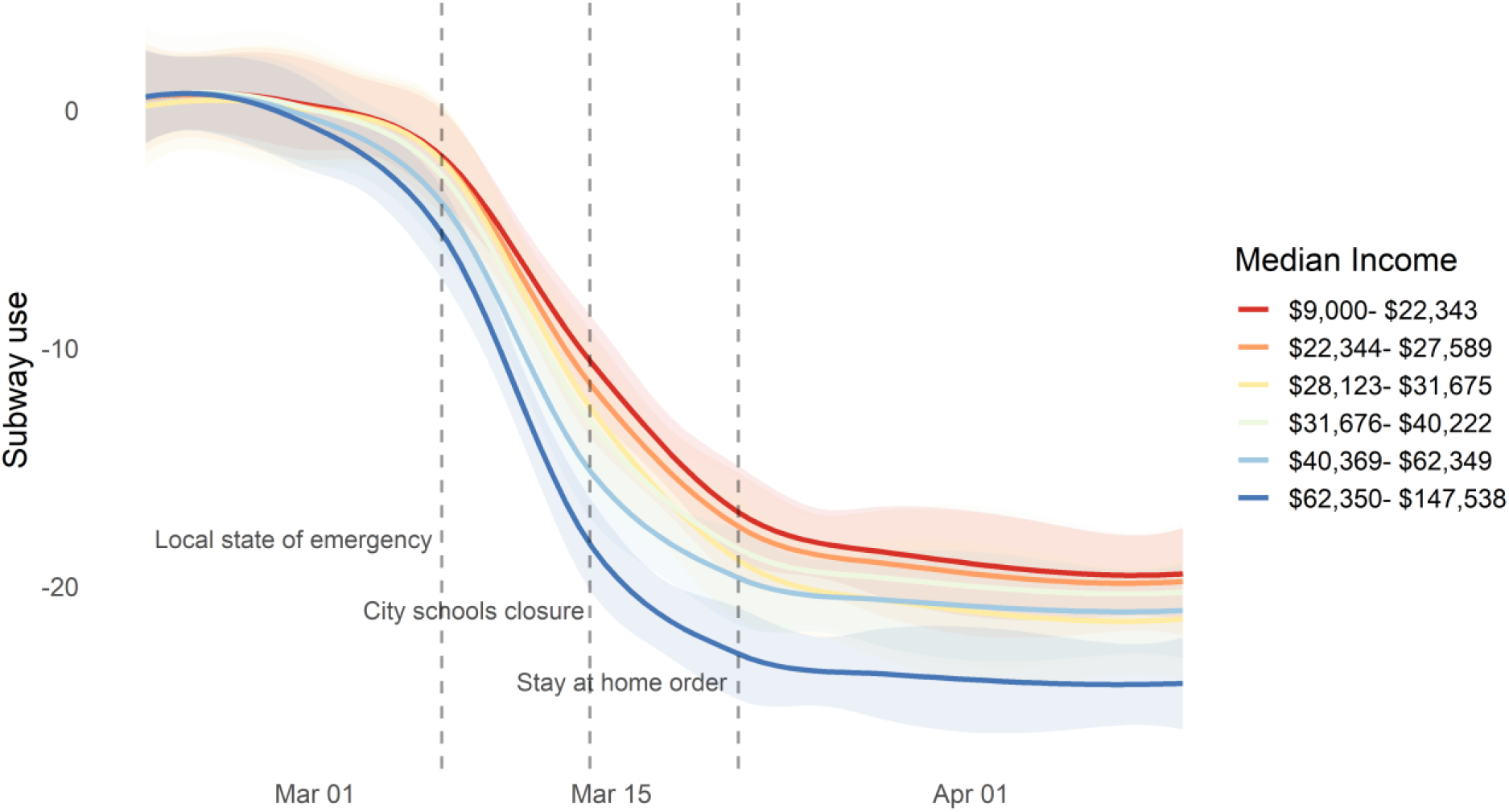
Change in subway use by median income sextiles. Loess smoothed lines and associated 95% confidence intervals were fitted over each income group. Vertical lines indicate timing of policies implemented in New York City.^38,54^

**Figure 3.**
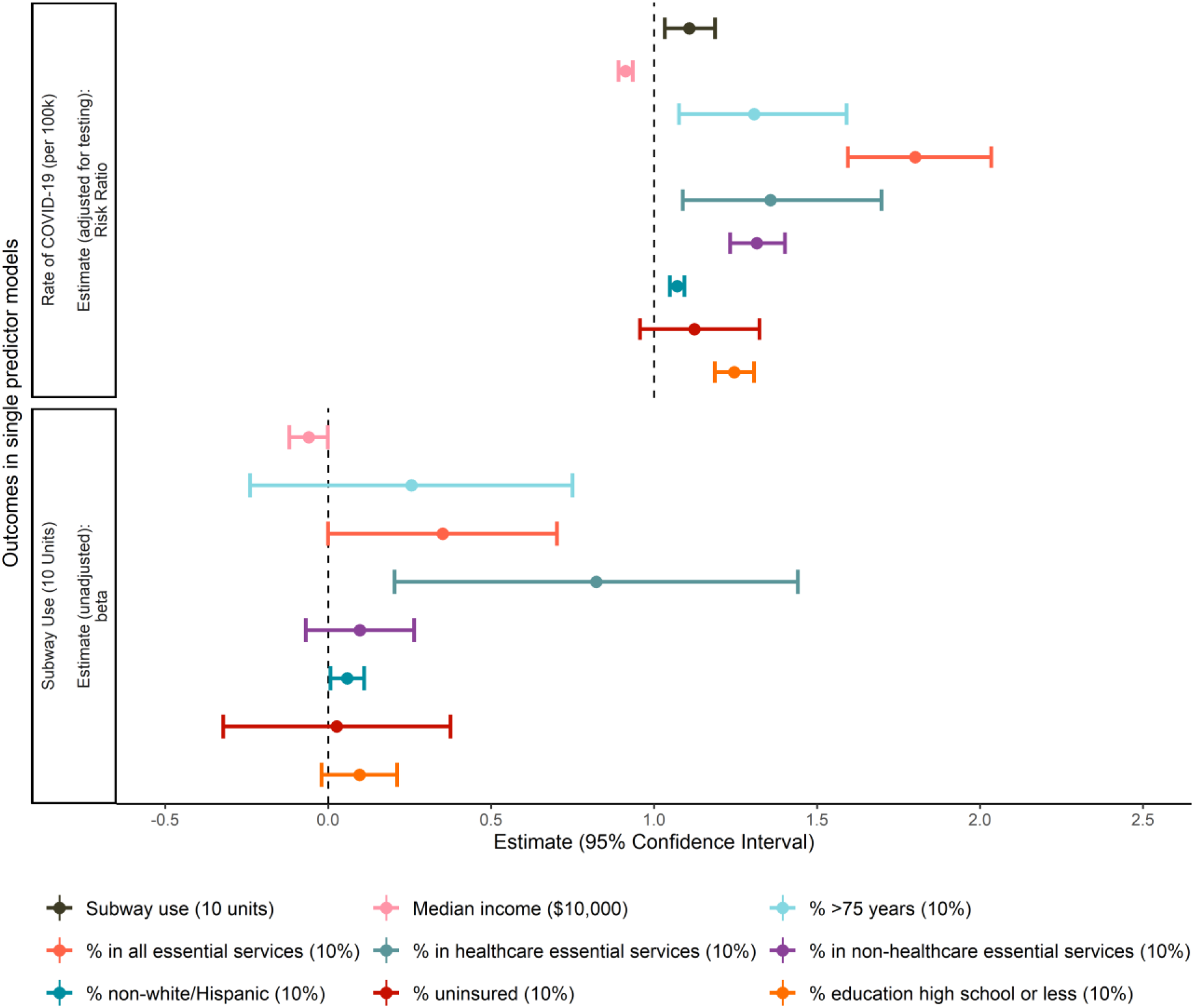
Associations between sociodemographic variables, subway use and COVID-19 rate per 100,000 population. All COVID-19 models were single predictor models adjusted for testing, to account for differential testing within ZCTAs. The subway outcomes are also from single predictor models (with no adjustments). The estimate for the rate of COVID-19 is a risk ratio with a null of 1, and the estimate for subway use is a slope (beta) with a null of 0. See associated supplementary Table 1 for more details.

### Subway use and COVID-19 outcomes

Smaller decreases in subway use during the pandemic period were significantly associated with the rate of positive cases per 100,000 population in each ZCTA population (RR = 1.12, 95% CI: 1.03–1.23). This association was slightly attenuated in analysis adjusted for testing (aRR = 1.11, 95% CI: 1.03–1.19), and decreased further when adjusted for testing and median income (aRR = 1.06, 95% CI: 1.00–1.12) (Figure 3, Supp Table 1). Smaller decreases in subway use only correlated with a greater proportion COVID-positive tests in the unadjusted analysis (RR = 1.03, 95% CI: 1.02–1.04); when adjusting for income, this association reversed, indicating that the proportion of COVID-cases among tested was negatively associated with subway use (aRR = 0.97, 95% CI: 0.96–0.98) (Supp Table 2). Smaller decreases in subway use were also associated with an increased rate of COVID-19 testing in both unadjusted (RR = 1.10, 95% CI: 1.03–1.18) and adjusted analysis (aRR = 1.07, 95% CI: 1.03–1.14) (Supp Table 2).

### Sociodemographic variables and COVID-19 outcomes

#### Rate of positive cases per 100,000 population

In analysis adjusting for the number of tests performed, all sociodemographic variables except percent uninsured were independently associated with the rate of positive COVID-19 cases per 100,000 population (Figure 3, Supp Table 1). An increase in median individual income of $10,000 was associated with a 10% decrease in the rate of COVID-19 (aRR = 0.90, 95% CI: 0.88–0.91), while an increase in 10% of the population in each ZCTA working in all essential services was associated with a nearly two-fold increase in the rate of positive cases per 100,000 population (aRR = 1.80, 95% CI: 1.59–2.04). A greater percent in healthcare essential services, greater percent in non-healthcare essential services, greater percent non-white/Hispanic, and greater percent education high school or less also increased the rate of positive COVID-19 cases per 100,000 population when adjusted for testing (Figure 3, Supp Table 1). When we adjust for both testing and median individual income, the associations remained; however, the percentage uninsured was associated with less reported COVID-19 (Supp Table 1).

Secondary COVID-19 outcomes (proportion positive among tested and rate of tests per 100,000 population)

All sociodemographic variables were associated with the proportion of positive tests in unadjusted analyses. The effect estimates of these exposures were generally in the same direction as the primary outcome of rate of positive cases, with the exception that the percent > 75 years old (OR = 0.69, 0.67–0.71) and percent in healthcare essential services (OR = 0.73, 95% CI: 0.70–0.75) were both protective (Supp Fig 3, Supp Table 2). When adjusted for median income, all associations remained significant, except for the percent employed in healthcare essential services. In unadjusted analysis, all sociodemographic variables, except the percent uninsured, were associated with the rate of tests per 100,000 population. Median individual income had an inverse relationship with rate of testing per 100,000 population, where an increase in $10,000 in median income corresponded with a 6% decrease in testing rates (RR = 0.94, 95% CI: 0.93–0.96). ZCTAs with larger percent of > 75 years old, greater percent in all essential services (including healthcare and non-healthcare), greater percent non-white/Hispanic, and greater percent education high school or less increased the rate of tests (Supp Fig 3, Supp Table 2). All associations remained when adjusted for median income, except for the percent non-white/Hispanic. In the adjusted model, an increase of 10% percent uninsured decreased the rate of COVID-19 tests by 21% (aRR = 0.79, 95% CI: 0.70–0.75) (Supp Table 2).

## BOROUGH ANALYSIS

### Segmented regression of change in subway use and COVID-19 outcomes

Segmented regression shows that the breakpoint, or change in subway use for all four boroughs, occurred within 6 days of each other. Subway use reduction was estimated to occur first in Manhattan starting on February 22, 2020 (95% CI: February 19 to February 24). This was followed by Brooklyn on February 24 (95% CI: February 21 to February 27), Queens on February 26 (95% CI: February 22 to February 29), and Bronx on February 27 (95% CI: February 24 to February 29) (Figure 4).

**Figure 4.**
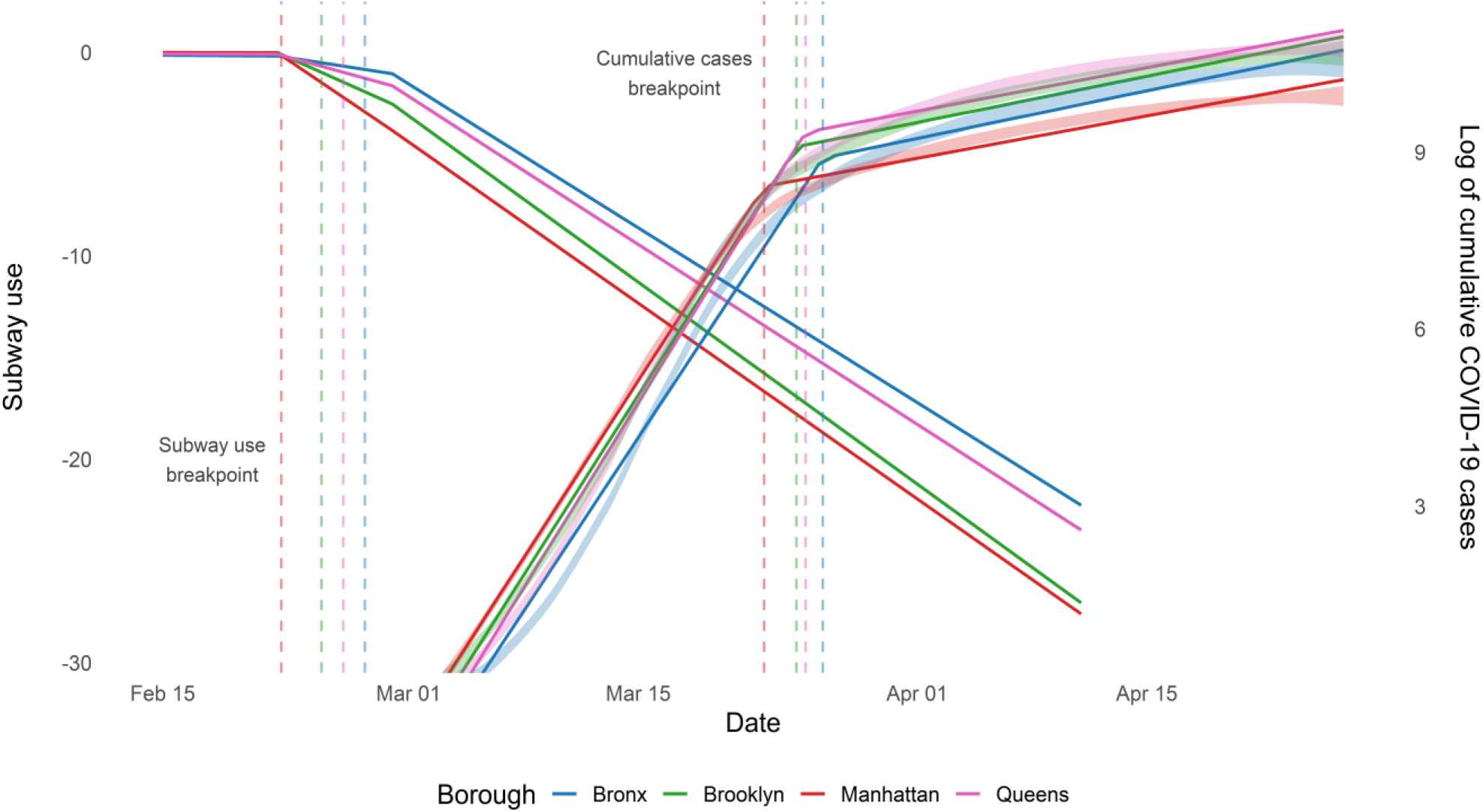
Segmented regression for subway use and log of cumulative cases, by borough. Opaque lines represent the fitted regression estimates, and transparent loess smoothed lines denote empirical case data. Vertical lines indicate the breakpoints of subway use (date of onset of decline in subway use) and of log of cumulative reported cases (end date of exponential growth period) for each borough.

There was a delay of approximately one month (mean 28.62 days) between the start of subway use reduction and end of the exponential growth period for reported cases (Figure 4). The end of the exponential growth period was estimated to end within a four-day period for four boroughs, with the Manhattan approximately stabilizing first on March 22 (95% CI: March 22 to March 23), Brooklyn on March 24 (95% CI: March 24 to March 25), Queens on March 25 (95% CI: March 24 to March 25), and the Bronx on March 26 (95% CI: March 22 to March 23). The date of decrease in subway use among the boroughs (Figure 5a) corresponded qualitatively to lower subway use on the week of April 11 (Figure 5b). These earlier decreases in subway use also had a similar trend to the exponential growth period, where boroughs that decreased subway use earlier experienced an earlier end of the exponential growth period (Figure 5c). The epidemic doubling time was the longest (slowest growth rate) for the Bronx, the borough with the last subway use decrease and the least amount of change in subway use (Figure 5d).

**Figure 5.**
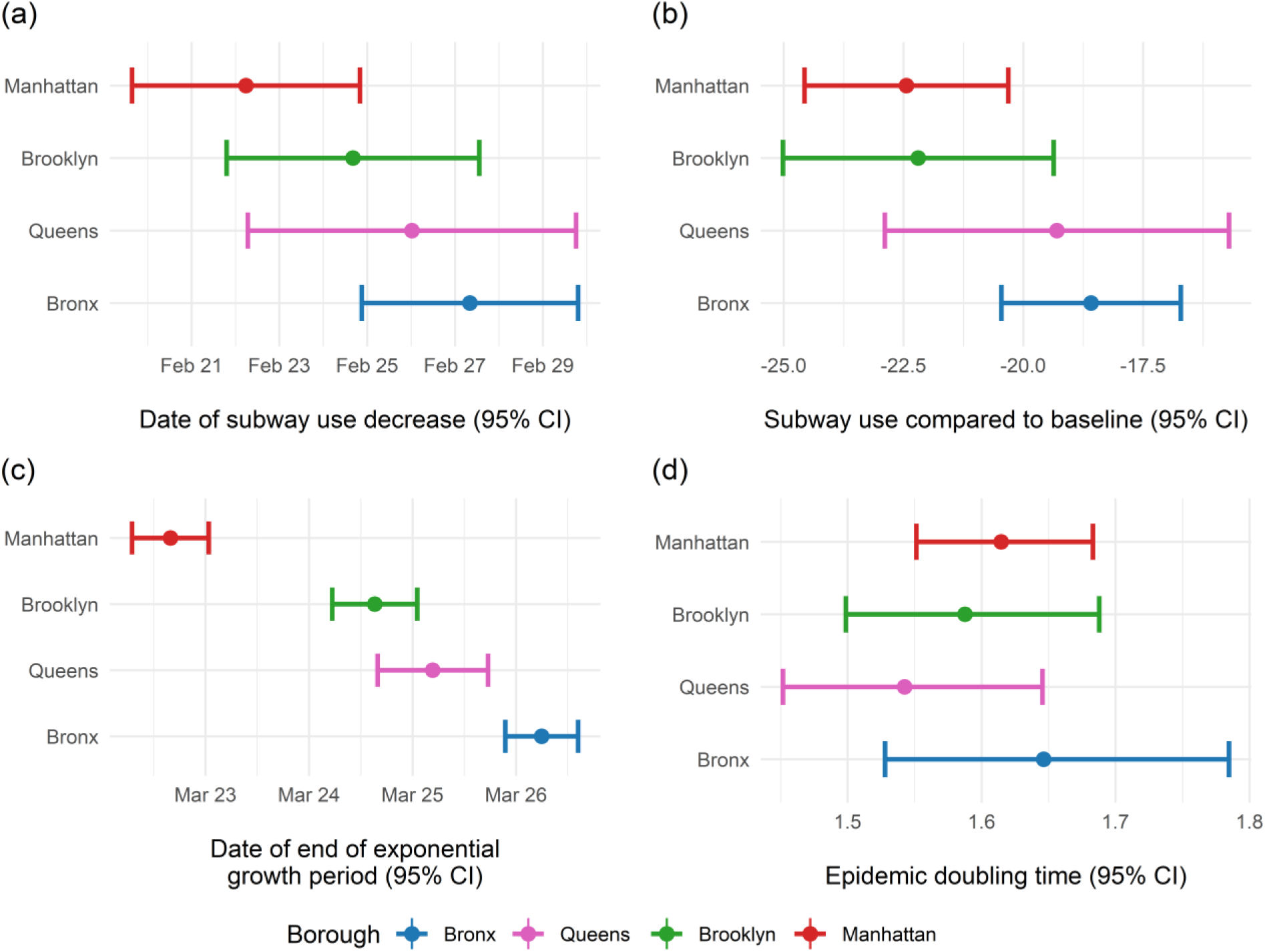
Subway use and epidemiological parameters of the four New York City boroughs. (a) Date of onset for the decline in subway use estimated by segmented regression; (b) Change in subway use compared to baseline; (c) End date of COVID-19 exponential growth period assessed through segmented regression; and (d) Epidemic doubling time based on rate of growth during the exponential growth period.

## DISCUSSION

On March 22, 2020, following the New York State on PAUSE executive order, a stay-at-home guidance was issued by New York City Mayor Bill de Blasio.^38^ However, we show that subway use started declining almost a full month prior to issuance and that the order’s timing roughly coincided with the end of the exponential growth period of COVID-19. While this suggests that many socially distanced on their own accord in response to the global reports of COVID-19, an **individual’s abilit**y to fully participate in non-pharmaceutical interventions varies across occupation, industry of employment, and socioeconomic status. Here we report evidence of substantial social distancing disparities throughout New York City. Communities with greater subway use were more likely to be socioeconomically disadvantaged, have a larger percentage of persons of color, and have a greater percentage of essential workers. Furthermore, these same communities were associated with greater COVID-19 burden, demonstrating inequities in both opportunity to reduce exposure and eventual COVID-19 infection. Areas with the lowest median individual income and a greater percentage of non-white and/or Hispanic/Latino individuals had smaller decreases in subway use, indicating that marginalized communities had reduced ability to shelter-in-place. However, these associations do not remain when adjusting for percent employed in essential services, suggesting that disparities in mobility reductions are driven by necessity, and consequently reduced privilege to socially distance. We also find that rates of SARS-CoV-2 testing is influenced by area-level sociodemographic variables, showing differential testing in New York City even when adjusted for median income.

It is widely established that social determinants of health lead to health inequities,^39–42^ and our findings are consistent with recent literature describing health disparities in COVID-19.^43–45^ Socioeconomic status is also associated with other underlying comorbidities that may heighten vulnerability to COVID-19 infection and mortality such as hypertension, obesity, renal disease, heart disease, and diabetes.^46–49^ Moreover, the ability to stay-at-home and physically distance are not only considerably more difficult in those engaged in essential work, but also in those with other adverse social determinants such as food insecurity,^17,18^ unstable housing,^19,20^ or experiencing domestic violence.^21,22^ These risk factors are further compounded by variability in testing access and volume across sociodemographic factors, which have been previously demonstrated across the United States^35,50^ and within New York City.^51^ The interrelationship of socioeconomic disparities, ability to social distance, chronic diseases, and COVID-19 is complex, but our findings demonstrate that the COVID-19 pandemic disproportionally affects the poorest and most vulnerable communities in New York City.

There are a number of limitations of this study. One limitation of the subway data is that a swipe represents an individual entering the subway station to take a trip, but the NYC subway, unlike some other underground systems, only require individuals to swipe when entering the subway; therefore, we do not know where trips terminated. Moreover, testing and reporting bias may distort the case counts for ZCTAs and boroughs. We attempted to limit the extent of this distortion by adjusting for tests given, but variability in volume is a function of both resource allocation and response to disease incidence and thus it is impossible to disentangle these biases. Additionally, mild and asymptomatic cases are likely underestimated for all of New York City and it is possible that our findings are related to differential ascertainment rather than true prevalence. However, our study found that income and race/ethnicity were predictors of COVID-19 prevalence in NYC, when adjusting for testing effort, and that lower-income neighborhoods and communities of color were hit hardest. This conclusion is supported by a recent report from **the NY State Governor’s office that stated** “*lower-income New York City communities and communities of color show 27 percent of individuals tested positive for COVID-19 antibodies, compared with 19.9 percent of New York City’s overall population*”.^52^ Further research is needed to clarify these disparities as COVID-19 seems to entrench existing inequalities and health disparities.

Our findings that social distancing inequities and health disparities are associated with SARS-CoV-2 infection are consistent with previous research in NYC.^51,53^ To our knowledge, this is one of the first studies to systematically assess the interrelationship between sociodemographic factors, mobility, and COVID-19. We show a 28-day lag time between the dramatic fall in subway ridership and the end of the exponential growth phase for reported cases of COVID-19, and heterogeneity in these reductions are associated with socioeconomic status. Our study provides further evidence that the most socially disadvantaged and poorest communities are not only at an increased risk for COVID-19 infection, but lack the privilege to fully engage in social distancing interventions, potentially compounding already existing health inequalities. COVID-19 is still a rapidly worsening crises, and the burden is increasing in many countries. In order to effectively fight this pandemic, sociodemographic and health disparities must be addressed.

## Data Availability

Weekly Metropolitan Transportation Authority (MTA) New York City transit subway data are publicly available. New York City Department of Health and Mental Hygiene COVID-19 data are available openly.

https://data.ny.gov/Transportation/Fare-Card-History-for-Metropolitan-Transportation-/v7qc-gwpn

https://github.com/nychealth/coronavirus-data

## SUPPLEMENT

**Supplementary Figure 1.**
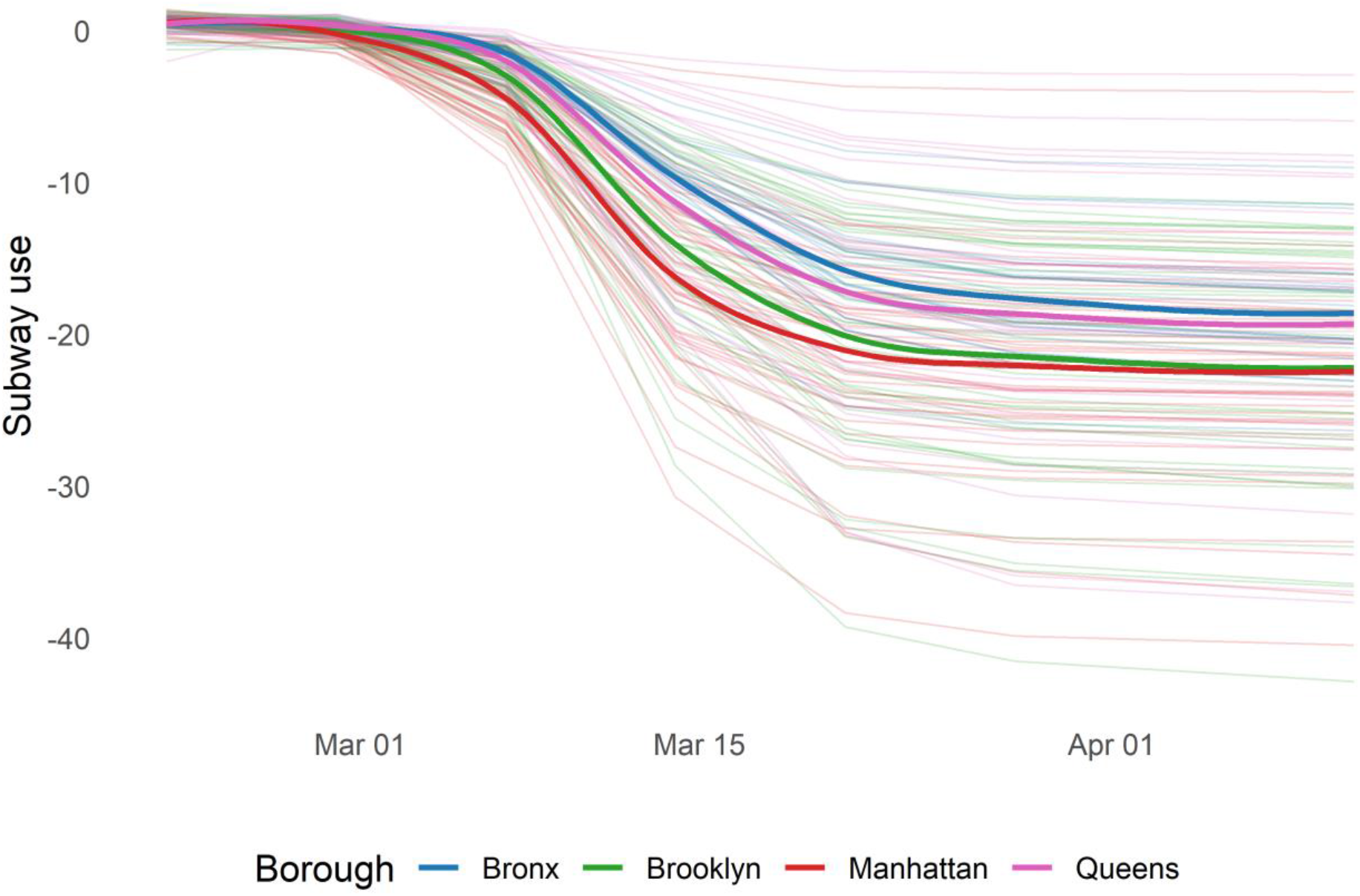
Relative change in subway use during the early SARS-CoV-2 outbreak (week of February 22 to April 11, 2020). Opaque lines represent loess fitted smoothed lines for the four boroughs, and the transparent lines denote the change in subway use among zip code tabulation areas; subway use change is calculated standardized to usage during January and February.

**Supplementary Figure 2.**
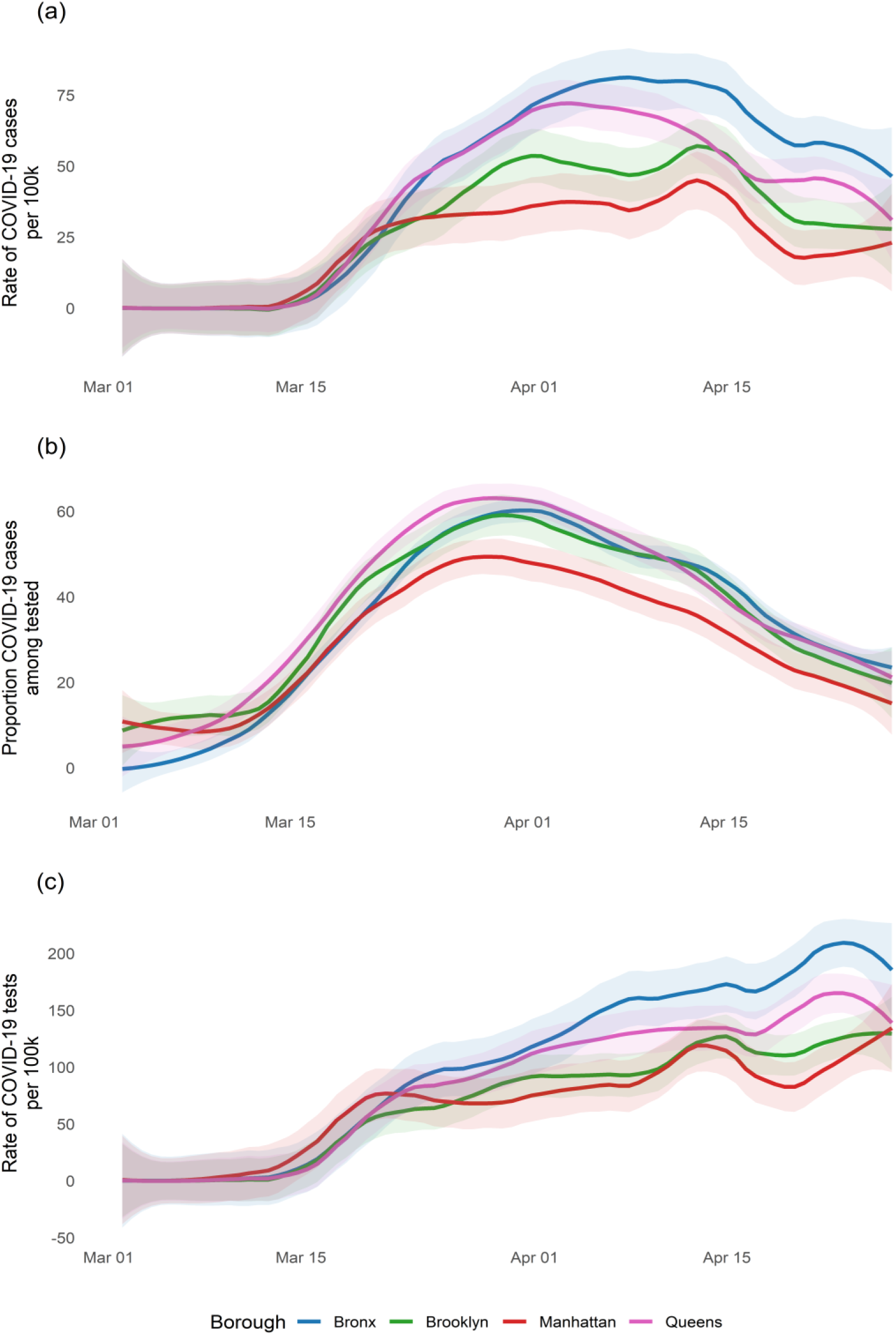
Loess smoothed line and associated 95% confidence intervals of (a) percent of positive cases among tested per 100,000 population; (b) rate of positive COVID-19 cases per 100,000 population; and (c) rate of COVID testing per 100,000 population, among boroughs.

**Supplementary Figure 3.**
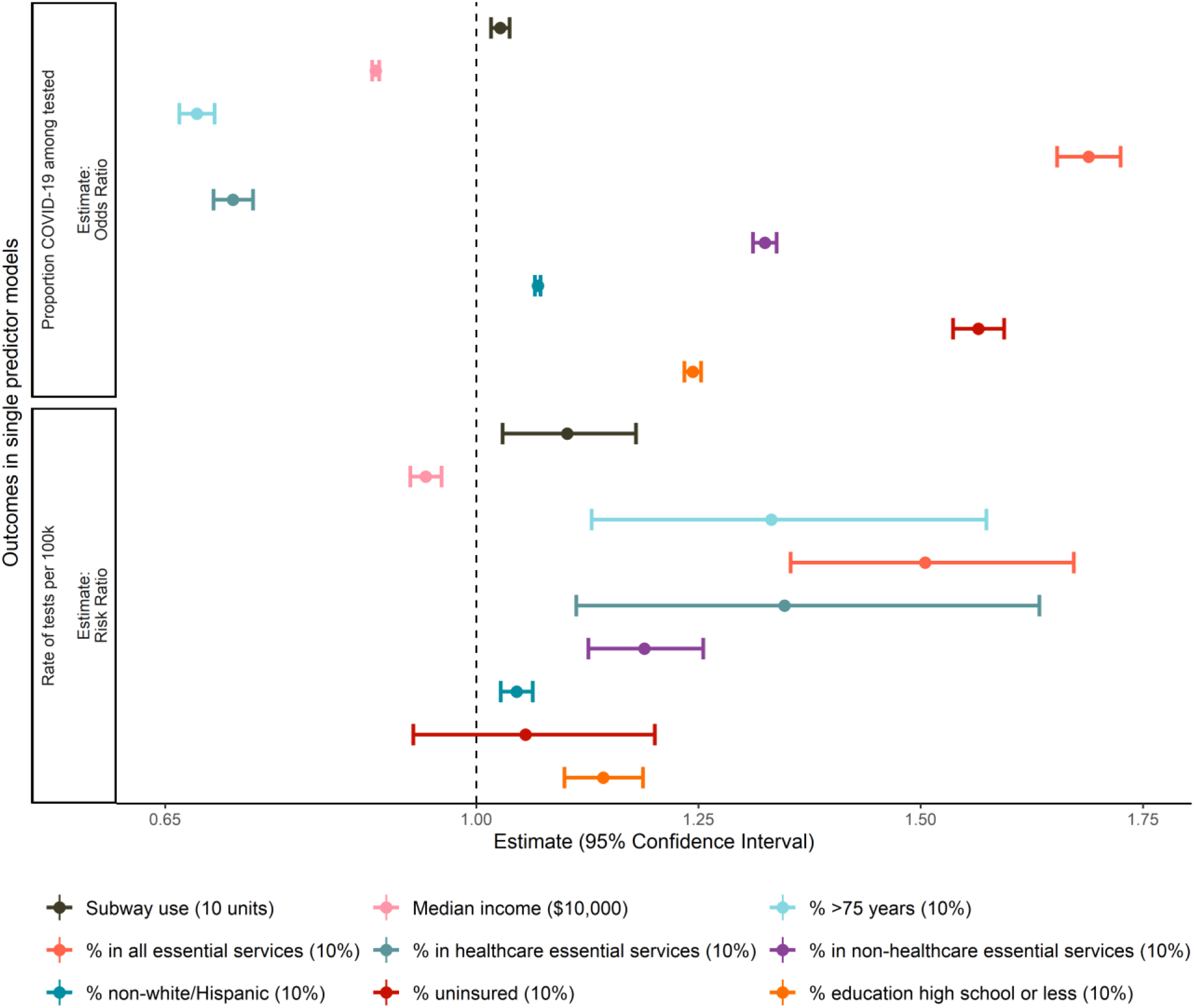
Unadjusted associations between sociodemographic variables, subway use change and proportion COVID-19 among tested and rate of tests per 100,000 population at the ZCTA-level. See associated supplementary Table 2 for more details.

**Supplementary Table 1.** Associations between sociodemographic variables, subway use and COVID-19 cases per 100,000 population. Each cell is one effect estimate for one model.

|  | Outcomes |  |  |  |  |
| --- | --- | --- | --- | --- | --- |
|  | Subway use<sup>i</sup> |  | Rate of positive cases per 100,000 population in the population<sup>ii</sup> |  |  |
|  | Unadjusted<br>*B* (95% CI) | Adjusted<br>*B* (95% CI)<sup>a</sup> | Unadjusted<br>RR (95% CI) | Adjusted<br>RR (95% CI)<sup>b</sup> | Adjusted<br>RR (95% CI)<sup>c</sup> |
| **Sociodemographic variables** |  |  |  |  |  |
| Median income (\$10,000) | **-0.06\***<br>(-0.12, -0.99) | -0.04<br>(-0.12, 0.05) | **0.90\*\*\*\***<br>(0.88, 0.91) | **0.91\*\*\*\***<br>(0.89, 0.93) | ----<sup>+</sup> |
| Percent >75 years (10%) | 0.26<br>(-0.24-0.75) | 0.27<br>(-0.22, 0.75) | 1.23<br>(0.98, 1.54) | **1.31\*\***<br>(1.08, 1.59) | **1.25\*\***<br>(1.06, 1.48) |
| Percent in essential services (10%) | **0.35\***<br>(0.01, 0.7) | ----<sup>+</sup> | **2.00\*\*\*\***<br>(1.76, 2.26) | **1.8\*\*\*\***<br>(1.59, 2.04) | **1.59\*\*\*\***<br>(1.36, 1.86) |
| Percent in healthcare essential services (10%) | **0.82\*\***<br>(0.2, 1.44) | ----<sup>+</sup> | 1.19<br>(0.93, 1.53) | **1.36\*\***<br>(1.09, 1.7) | **1.46\*\***<br>(1.21, 1.77) |
| Percent in non-healthcare essential services (10%) | 0.10<br>(-0.07, 0.26) | ----<sup>+</sup> | **1.39\*\*\*\***<br>(1.3, 1.48) | **1.31\*\*\*\***<br>(1.23, 1.4) | **1.22\*\*\*\***<br>(1.11, 1.33) |
| Percent non-white/Hispanic (10%) | **0.06\***<br>(0.01, 0.11) | 0.04<br>(-0.02, 0.1) | **1.09\*\*\*\***<br>(1.07, 1.11) | **1.07\***<br>(1.05, 1.09) | **1.04\*\*\*\***<br>(1.01, 1.06) |
| Percent with uninsured (10%) | 0.03<br>(-0.32, 0.38) | -0.29<br>(-0.72, 0.14) | **1.34\*\*\***<br>(1.14, 1.58) | 1.12<br>(0.96, 1.32) | **0.84\*\***<br>(0.73, 0.98) |
| Percent education high school or less (10%) | 0.10<br>(-0.02, 0.21) | 0.01<br>(-0.18, 0.2) | **1.29\*\*\*\***<br>(1.23, 1.35) | **1.25\*\*\*\***<br>(1.19, 1.31) | **1.22\*\*\*\***<br>(1.11, 1.34) |
| **Human mobility** |  |  |  |  |  |
| Subway use change (10 units) | ----<sup>+</sup> | ----<sup>+</sup> | **1.12\*\*\*\***<br>(1.03, 1.23) | **1.11\*\***<br>(1.03, 1.19) | 1.06<br>(1.00, 1.12) |

pvalue \* < 0.05; \*\* < 0.01; \*\*\* < 0.001; \*\*\*\* < 0.0001
<sup>+</sup> No adjusted estimate
<sup>i</sup> Linear model
<sup>ii</sup> Generalized linear model with a negative binomial distribution and a log link, total population as offset
<sup>a</sup> Adjusted for percent employed in essential services
<sup>b</sup> Adjusted for number of tests
<sup>c</sup> Adjusted for number of tests and median income

**Supplementary Table 2.**
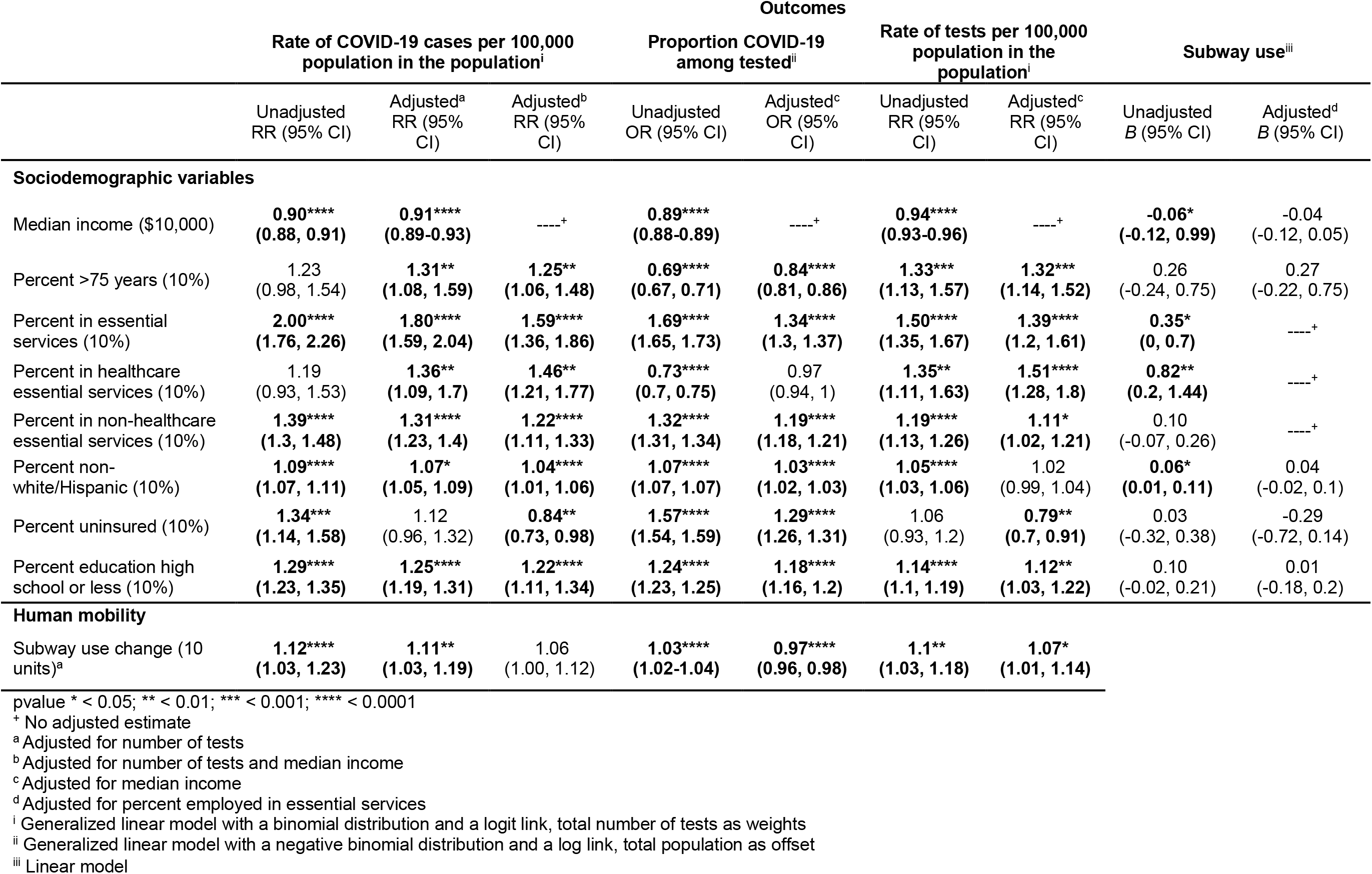
Associations between sociodemographic variables, subway use change and all COVID-19 outcomes at the ZCTA-level.

## Author contributions

K. Sy, M. Martinez, L. White contributed to conceptualization. K. Sy and M. Martinez contributed to data acquisition. K. Sy and B. Rader contributed to data analysis. All authors contributed to interpretation of results and manuscript writing.

## Acknowledgements

The authors would like to thank Kathryn Cordiano and Hanbin Wang Jr. for their thoughtful contribution to the manuscript.

